# Development and External Validation of a Diagnostic Model for Intra-Procedural Hypotension during Primary Percutaneous Coronary Intervention

**DOI:** 10.1101/2020.06.15.20132209

**Authors:** Yong Li

## Abstract

**Background:** Intra-procedural hypotension weaken the benefit of primary percutaneous coronary intervention (PPCI) and worsens the prognosis of acute ST elevation myocardial infarction (STEMI) patients.

**Objectives:** To develop and externally validate a diagnostic model of intra-procedural hypotension.

**Methods:** Design:Multivariable logistic regression of a cohort of acute STEMI patients. Setting: Emergency department ward of a university hospital. Participants: Diagnostic model development: Totally 1239 acute STEMI patients who were consecutively treated with PPCI from November 2007 to December 2013. External validation: Totally 1294 acute STEMI patients who were treated with PPCI from January 2014 to June 2018. Outcomes: Intra-procedural hypotension. Intra-procedural hypotension was defined as pre-procedural systolic blood pressure (SBP) was > 90mmHg, intra-procedural SBP less than or equal to 90 mmHg persistent or transient.

**Results:** Totally 121(9.8%) patients presented intra-procedural hypotension in the development dataset; 123 (9.5%) patients presented intra-procedural hypotension in the validation dataset. The strongest predictors of intra-procedural hypotension were no-reflow, the culprit vessel was not left anterior descending, complete occlusion of culprit vessel, using thrombus aspiration devices during operation, and without history of diabetes. We developed a diagnostic model of intra-procedural hypotension. The area under the receiver operating characteristic (ROC) curve (AUC) was 0.685 ± 0.022, 95% CI = 0.641 ~ 0.729 in the development set. We constructed a nomogram using the development database based on predictors of intra-procedural hypotension. The AUC was 0.718 ±0.022, 95% CI = 0.674 ~ 0.761 in the validation set. Discrimination, calibration, and decision curve analysis were satisfactory.

**Conclusions:** We developed and externally validated a diagnostic model of intra-procedural hypotension during PPCI. We can use the formula or nomogram to predict intra-procedural hypotension.

This study was registered with WHO International Clinical Trials Registry Platform (ICTRP) on 6 September 2019 (registration number:ChiCTR1900025706). http://www.chictr.org.cn/edit.aspx?pid=42913&htm=4.

## Introduction

Primary percutaneous coronary intervention(PPCI) is the best available reperfusion strategy for ST elevation myocardial infarction (STEMI). Intra-procedural hypotension weaken the benefit of PPCI and worsens the prognosis of patients. ^[1–3]^ We want to develop and externally validate a diagnostic model of intra-procedural hypotension. The aim of our study was 4-fold: (1) to identify predictive factors; (2) to develop a diagnostic model; (3) to construct a nomogram; and (4) to externally validate diagnostic model.

## Methods

We used type 2b of prediction model studies covered by Transparent Reporting of a multivariable prediction model for Individual Prognosis Or Diagnosis (TRIPOD) statement. ^[4]^We split the data nonrandomly by time into 2 groups: one to develop the prediction model and one to evaluate its predictive performance. ^[4]^Type 2b was referred to as “external validation studies”. ^[4]^

The derivation cohort was 1239 patients with acute STEMI presenting within 12 hours from the symptom onset who were consecutively treated with PPCI between November 2007 and December 2013 in Beijing Anzhen Hospital, Capital Medical University.

The validation cohort was 1294 patients we recruited on the same basis between January 2014 and June 2018 in Beijing Anzhen Hospital, Capital Medical University.

Inclusion criteria: STEMI patients presenting within 12 hours from the symptom onset who were treated with PPCI. We established the diagnosis of acute myocardial infarction (AMI) and STEMI base on the fourth universal definition of myocardial infarction. ^[5]^

Exclusion criteria: 1. patients received thrombolysis; 2. patients received bivalirudin.

Since we performed retrospective analysis, the requirement of informed consent was waived by Ethics Committee of Beijing Anzhen Hospital Capital Medical University.

Intra-procedural hypotension was defined as pre-procedural systolic blood pressure (SBP) was > 90mmHg, intra-procedural SBP less than or equal to 90 mmHg persistent or transient. ^[6]^ Outcome of interest was intra-procedural hypotension.

The presence or absence of intra-procedural hypotension was decided blinded to the predictor variables, based on consensus, and recorded on the operation record.

We selected 9 predictor variables for inclusion in our prediction rule from the larger set according to clinical relevance and the results of baseline descriptive statistics in our cohort of pre-experiment.

They were shown in Table 1.

**Table 1.**
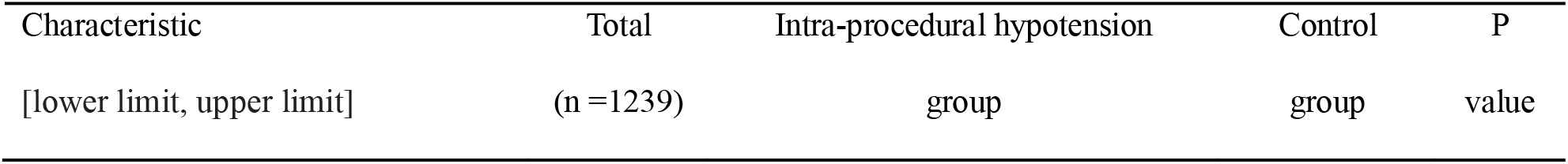
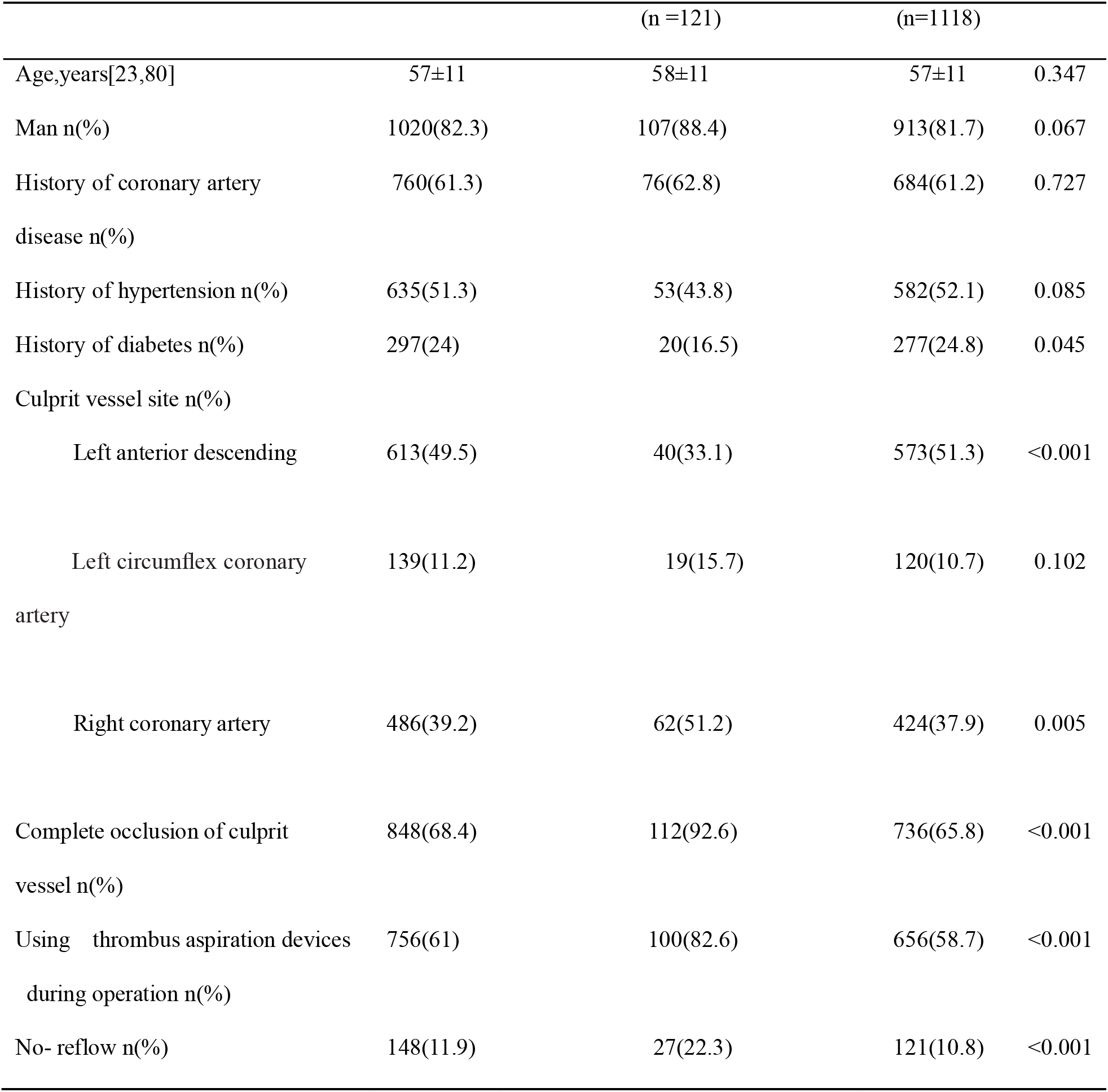
Demographic, clinical, and angiographic characteristics of patients with intra-procedural hypotension and control during PPCI in the development data sets

| Characteristic | Total | Intra-procedural hypotension | Control | P |
| --- | --- | --- | --- | --- |
| [lower limit, upper limit] | (n =1239) | group | group | value |

|  |  | (n =121) | (n=1118) |  |
| --- | --- | --- | --- | --- |
| Age,years[23,80] | 57±11 | 58±11 | 57±11 | 0.347 |
| Man n(%) | 1020(82.3) | 107(88.4) | 913(81.7) | 0.067 |
| History of coronary artery disease n(%) | 760(61.3) | 76(62.8) | 684(61.2) | 0.727 |
| History of hypertension n(%) | 635(51.3) | 53(43.8) | 582(52.1) | 0.085 |
| History of diabetes n(%) | 297(24) | 20(16.5) | 277(24.8) | 0.045 |
| Culprit vessel site n(%) |  |  |  |  |
| Left anterior descending | 613(49.5) | 40(33.1) | 573(51.3) | <0.001 |
| Left circumflex coronary artery | 139(11.2) | 19(15.7) | 120(10.7) | 0.102 |
| Right coronary artery | 486(39.2) | 62(51.2) | 424(37.9) | 0.005 |
| Complete occlusion of culprit vessel n(%) | 848(68.4) | 112(92.6) | 736(65.8) | <0.001 |
| Using thrombus aspiration devices during operation n(%) | 756(61) | 100(82.6) | 656(58.7) | <0.001 |
| No- reflow n(%) | 148(11.9) | 27(22.3) | 121(10.8) | <0.001 |

We used angiographic criteria for the diagnosis of no-reflow. ^[7]^ No-reflow was defined as Thrombolysis In Myocardial Infarction flow grade (TIMI)<3. ^[8]^

Some people suggested that each candidate variable must have at least 10 events for model derivation and at least 100 events for model validation.^[4]^ Our number of samples and events exceeded expectations, so it was expected to provide very reliable estimates.^[4]^

To ensure reliability of data, we excluded patients who had missing information on key predictors: no-reflow, the culprit vessel was left anterior descending (LAD), complete occlusion of culprit vessel, using thrombus aspiration devices during operation, and history of diabetes.

### Statistical Analysi

All continuous data had kept as continuous and retained on the original scale. We used multivariable logistic regression models to identify the correlates of intra-procedural hypotension during PPCI. We constructed a multivariable logistic regression model using the backward variable selection method. We used the Akaike information criterion (AIC)and Bayesian information criterion(BIC)to select predictors; it accounts for model fit while penalizing for the number of parameters being estimated and corresponds to using α = 0.157. ^[4]^

We assessed the predictive performance of the diagnostic model in the validation data sets by examining measures of discrimination, calibration, and decision curve analysis (DCA). ^[4]^

We performed statistical analyses with STATA version 15.1 (StataCorp, College Station, TX), R version 4.0.0(R Development Core Team; http://www.r-project.org), and the RMS package developed by Harrell (Harrell et al). All tests were two-sided and a P value <0.05 was considered statistically significant.

## Results

We drew a flow diagrams (Figure 1).

**Figure 1.**
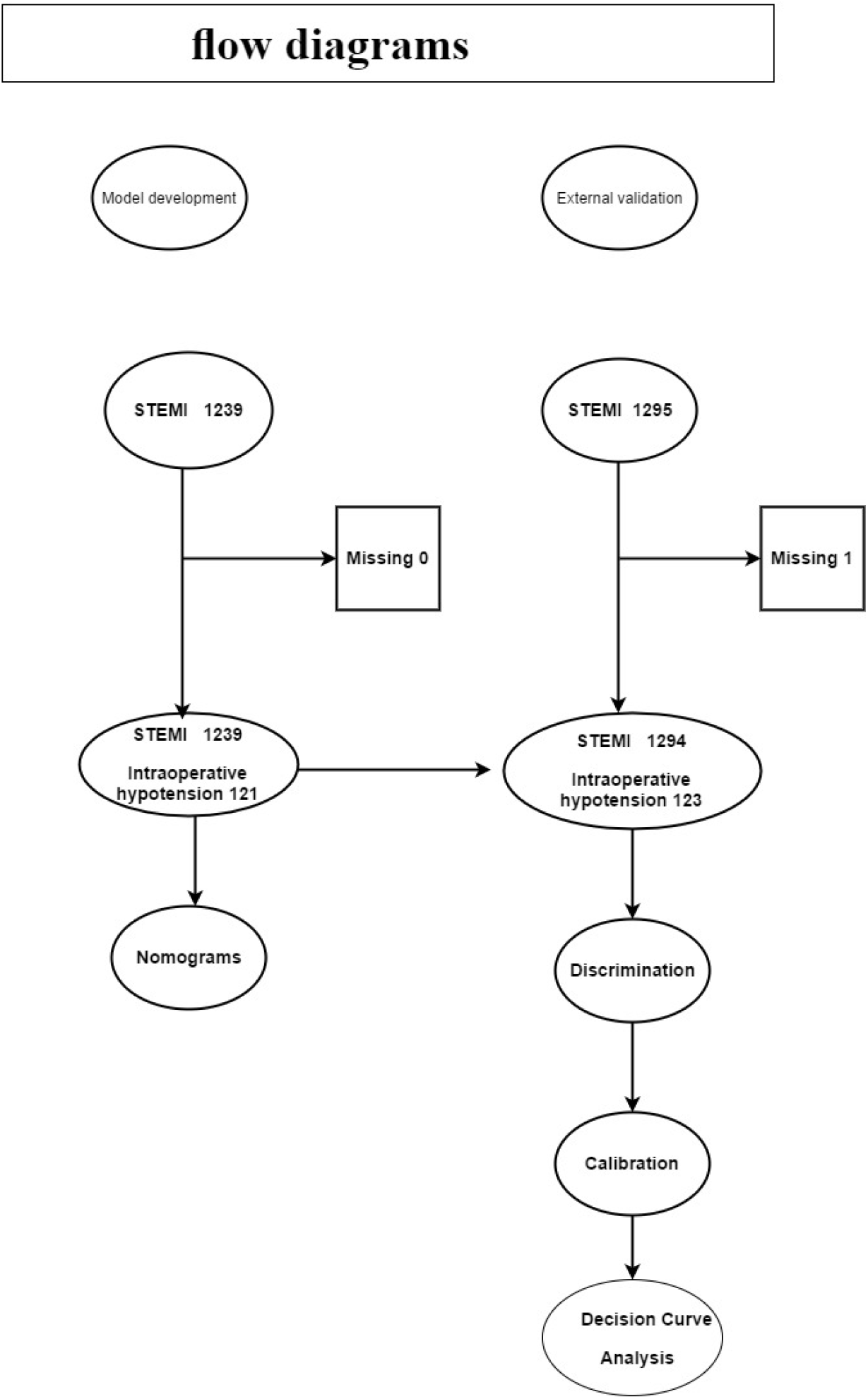
Flow diagrams.

Totally 121 patients had intra-procedural hypotension (intra-procedural hypotension group) and 1118 patients had no intra-procedural hypotension (control group) in the development data set. Baseline characteristics of the patients was shown in Table1.

After application of backward variable selection method, AIC, and BIC, five variables (no-reflow, the culprit vessel was LAD, complete occlusion of culprit vessel, using thrombus aspiration devices during operation, and history of diabetes) remained as significant independent predictors of intra-procedural hypotension. Odds ratio and coef were shown in Table 2 and Table 3.

**Table 2.** **Predictor of intra-procedural hypotension obtained from multivariable logistic regression models in the development data sets(odds ratio**)

| Intra-procedural hypotension | Odds ratio | Std.Err | Z | P>\|Z\| | 95% CI |
| --- | --- | --- | --- | --- | --- |
| The culprit vessel was LAD | .488 | .101 | -3.47 | 0.001 | .326 ~ .732 |
| Using thrombus aspiration devices during operation | 1.793 | .483 | 2.17 | 0.030 | 1.058 ~ 3.039 |
| No-reflow | 1.911 | .472 | 2.62 | 0.009 | 1.177 ~ 3.102 |
| History of diabetes | .589 | .154 | -2.02 | 0.043 | .353~.983 |
| Complete occlusion of culprit vessel | 4.351 | 1.643 | 3.90 | <0.001 | 2.076 ~ 9.12 |
| _cons | .0289 | .011 | -9.72 | <0.001 | .0141 ~ .059 |

**Table 3.** **Predictor of intra-procedural hypotension obtained from multivariable logistic regression models in the development data sets(coef)**

| Intra-procedural hypotension | Coef | Std.Err | Z | P>\| Z \| | 95% CI |
| --- | --- | --- | --- | --- | --- |
| The culprit vessel was LAD | -.717 | .207 | -3.47 | 0.001 | -1.122 ~ -.312 |
| Using thrombus aspiration devices during operation | .584 | .269 | 2.17 | 0.030 | .0560 ~ 1.112 |
| No-reflow | .648 | .247 | 2.62 | 0.009 | .163 ~ 1.132 |
| History of diabetes | -.529 | .261 | -2.02 | 0.043 | -1.042 ~ -.0167 |
| Complete occlusion of culprit vessel | 1.470 | .378 | 3.90 | <0.001 | .731 ~ 2.210 |
| _cons | -3.544 | .365 | -9.72 | <0.001 | -4.259 ~ -2.830 |

According to the above risk factors,we can calculate the predicted probability of intra-procedural hypotension using the following formula: P = 1/(1+exp(-(−3.544 +1.470* COCV +.648* CNR +-.529* HD+ −.717 * LAD + .584* TA))). COCV = complete occlusion of culprit vessel(0=No, 1=Yes); CNR = coronary artery no-reflow phenomenon(0=No, 1=Yes); HD = history of diabetes(0=No, 1=Yes); LAD = the culprit vessel was LAD(0=No, 1=Yes); TA = using thrombus aspiration devices during operation (0=No, 1=Yes).

We drew the ROC curve. AUC was 0.685 ± 0.022, 95% CI = 0.641 ~ 0.729.

We constructed the nomogram(Figure 2)using the development database based on the five independent predictors: No-reflow, the culprit vessel was LAD, complete occlusion of culprit vessel, using thrombus aspiration devices during operation, and history of diabetes.

**Figure 2.**
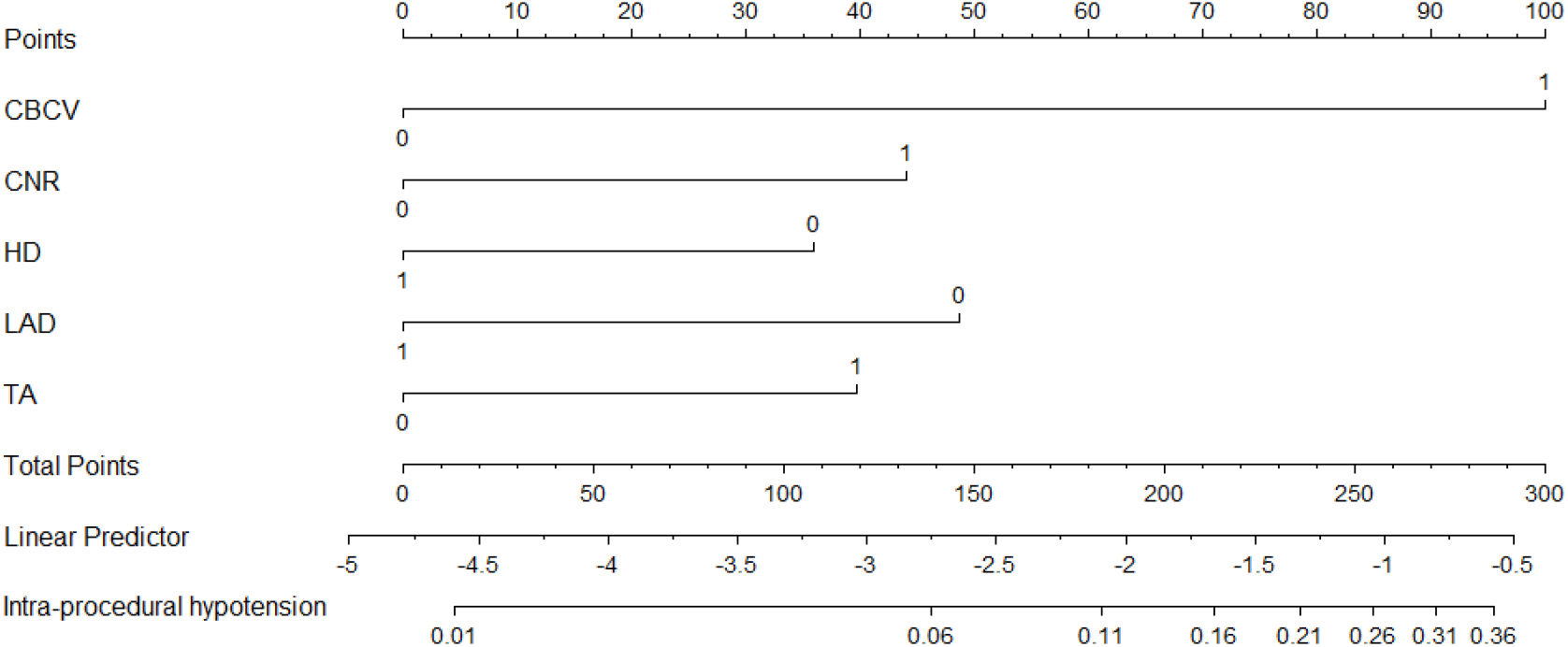
A nomogram for predicting intra-procedural hypotension during PPCI in patients with acute STEMI. CBCV = complete occlusion of culprit vessel(0=No, 1=Yes); CNR = coronary artery no-reflow phenomenon(0=No, 1=Yes); HD = history of diabetes(0=No, 1=Yes); LAD = the culprit vessel was LAD(0=No, 1=Yes); TA = using thrombus aspiration devices during operation(0=No, 1=Yes).

Totally 1294 acute STEMI patients in the validation data sets. Baseline characteristics of the patients was shown in Table 4.

**Table 4.**
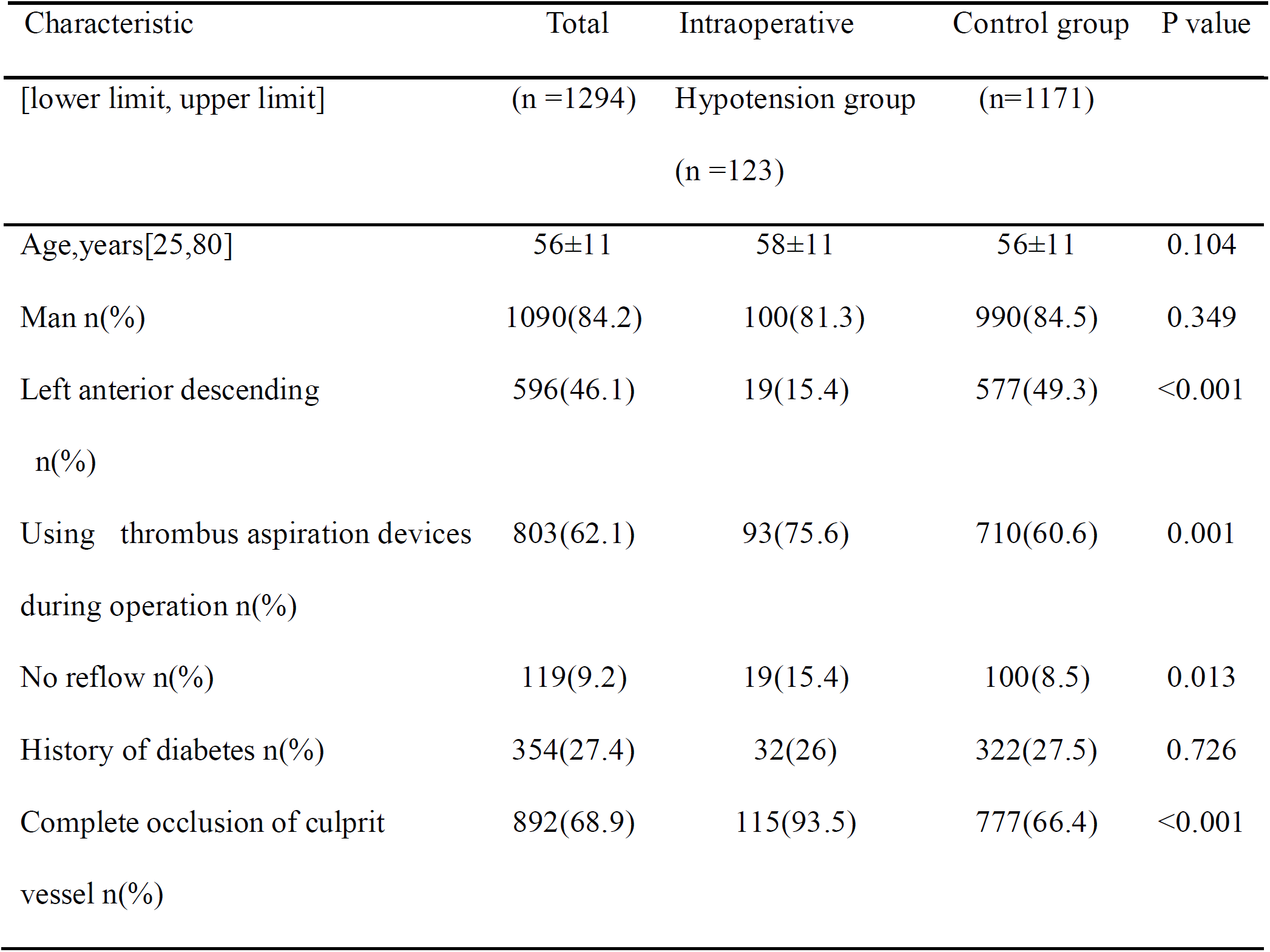
**Demographic,clinical,and angiographic characteristics of patients with intra-procedural hypotension and control group during PPCI in the validation data sets**

| Characteristic | Total | Intraoperative | Control group | P value |
| --- | --- | --- | --- | --- |

| [lower limit, upper limit] | (n =1294) | Hypotension group<br>(n =123) | (n=1171) |  |
| --- | --- | --- | --- | --- |
| Age,years[25,80] | 56±11 | 58±11 | 56±11 | 0.104 |
| Man n(%) | 1090(84.2) | 100(81.3) | 990(84.5) | 0.349 |
| Left anterior descending<br>n(%) | 596(46.1) | 19(15.4) | 577(49.3) | <0.001 |
| Using thrombus aspiration devices<br>during operation n(%) | 803(62.1) | 93(75.6) | 710(60.6) | 0.001 |
| No reflow n(%) | 119(9.2) | 19(15.4) | 100(8.5) | 0.013 |
| History of diabetes n(%) | 354(27.4) | 32(26) | 322(27.5) | 0.726 |
| Complete occlusion of culprit<br>vessel n(%) | 892(68.9) | 115(93.5) | 777(66.4) | <0.001 |

We can calculate the predicted probability of intra-procedural hypotension using the following formula: P = 1/(1+exp(-(−3.544 +1.470* COCV +.648* CNR +-.529*HD+ −.717 * LAD + .584* TA))). COCV = complete occlusion of culprit vessel(0=No, 1=Yes); CNR = coronary artery no-reflow phenomenon(0=No, 1=Yes); HD = history of diabetes(0=No, 1=Yes); LAD = the culprit vessel was LAD(0=No, 1=Yes); TA = using thrombus aspiration devices during operation (0=No, 1=Yes).

We drew the ROC curve. AUC was 0.718 ±0.022, 95% CI = 0.674 ~ 0.761.

We drew a calibration plot(Figure 3)with distribution of the predicted probabilities for individuals with and without the intra-procedural hypotension in the evalidation data sets.Hosmer-Lemeshow chi2(10) = 6.92, Prob > chi2 = 0.545 >0.05. Brier score = 0.0819< 0.25.

**Figure 3.**
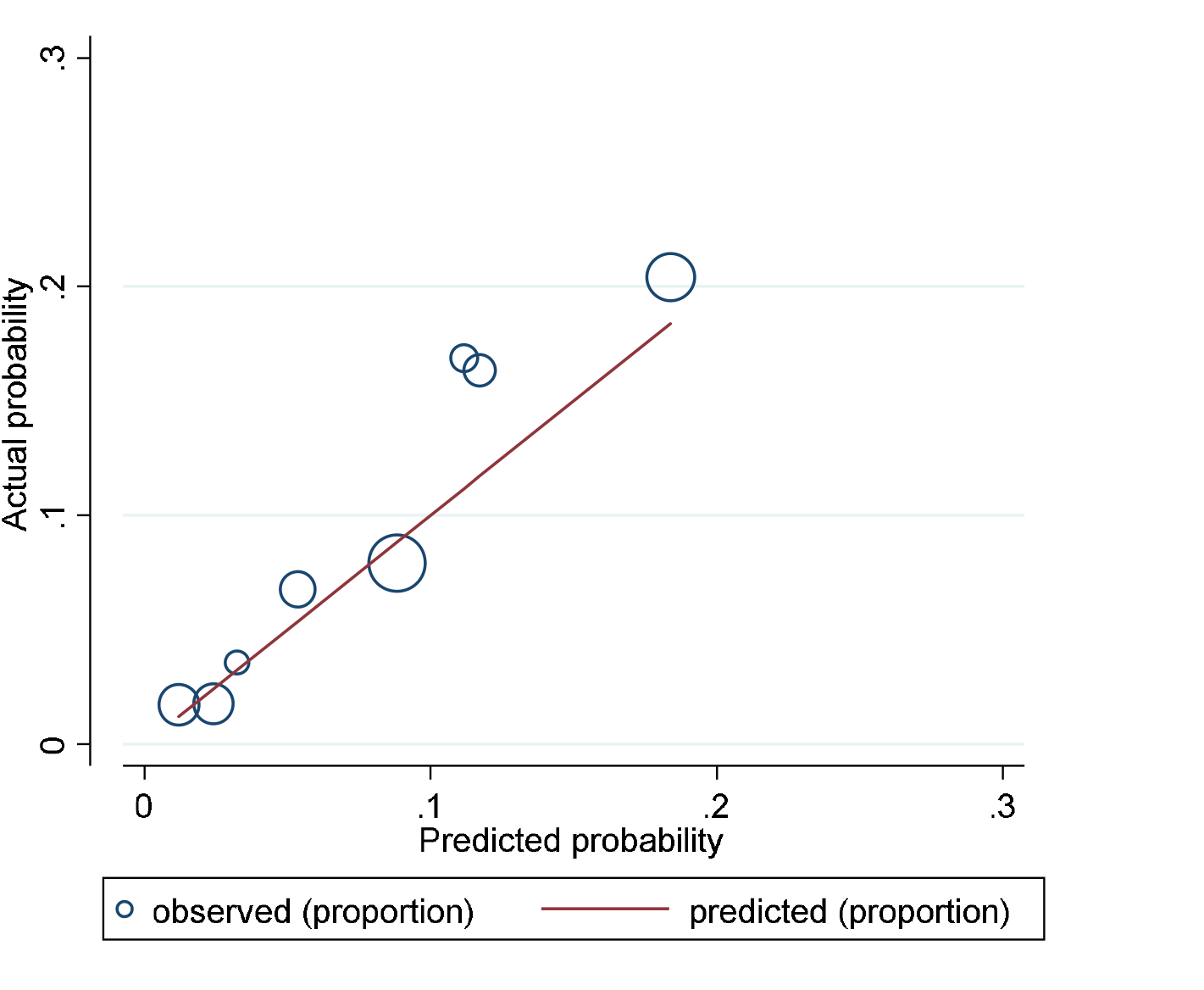
A calibration plot with distribution of the predicted probabilities for individuals with and without intra-procedural hypotension in the evalidation data sets.

DCA(Figure 4)in the validation data sets.

**Figure 4.**
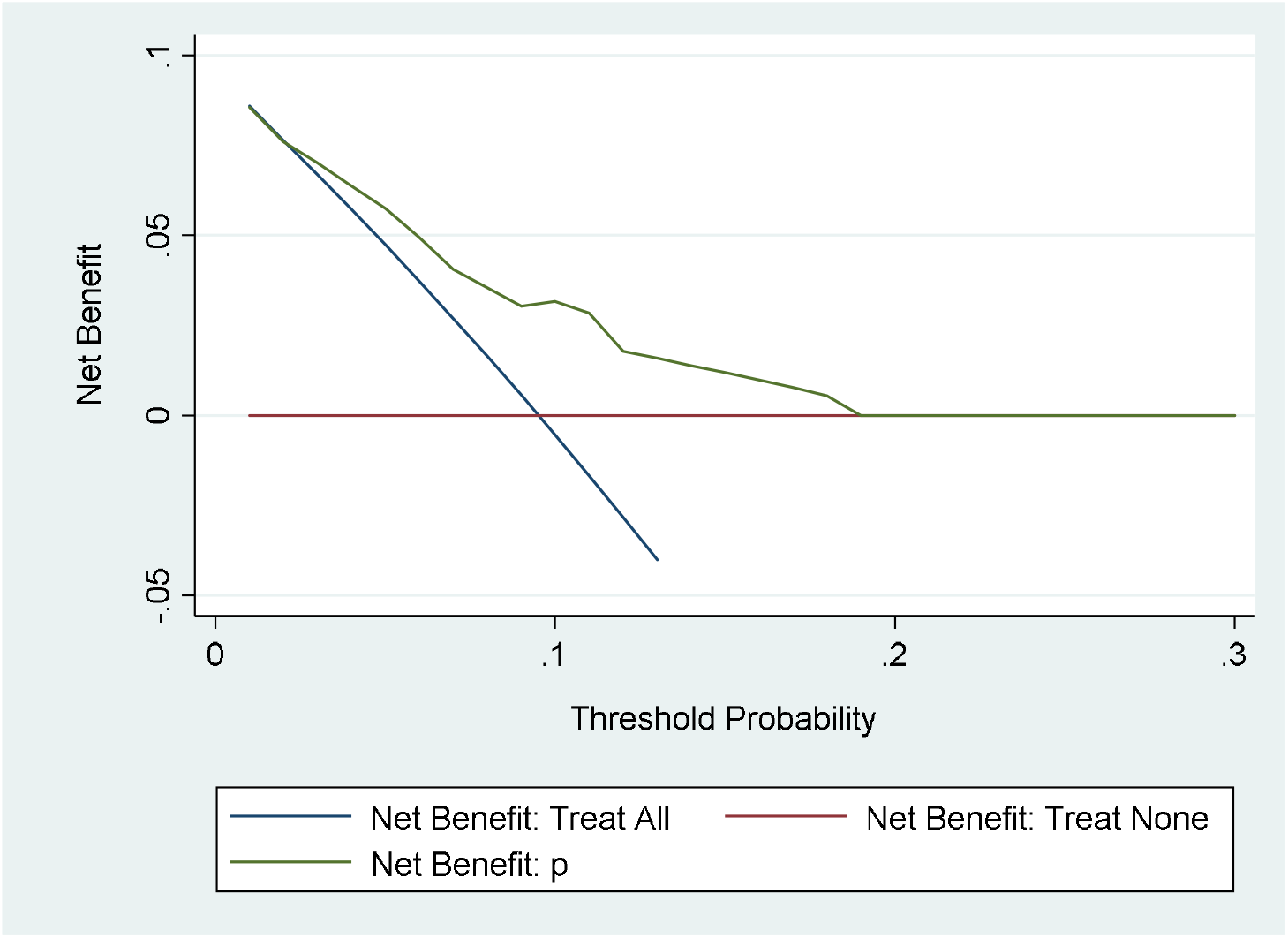
DCA in the evalidation data sets.

## Discussion

In this study, we investigated the predisposing factors of intra-procedural hypotension in patients with STEMI undergoing PPCI. A frequency of intra-procedural hypotension was 9.7% (121/1239) in the development data sets. No-reflow, the culprit vessel was not LAD, complete occlusion of culprit vessel, using thrombus aspiration devices during operation, and without history of diabetes were independent risk factors predicting intra-procedural hypotension. AUC was 0.718 ±0.022, 95% CI = 0.674 ~ 0.761 in the validation data sets. Prob > chi2 = 0.545 >0.05. Brier score = 0.0819< 0.25. Discrimination, calibration, and DCA were satisfactory. We can use the formula or nomogram to predict intra-procedural hypotension. This may enable patients at heightened risk of intra-procedural hypotension to be treated with the most appropriate individualised treatment early.

The culprit vessel is LAD is less likely to evoke intra-procedural hypotension. RCA or Left circumflex coronary artery(LCX) supply blood to inferior wall which preferential distribution of vagal nerve. No-reflow is associated with intra-procedural hypotension. Myocardial reperfusion can trigger excitement of the heart’s vagal nerve endings, ^[9]^ which can cause coronary artery spasm and elicit no-reflow.^[10]^ The excitement of the vagal nerve is called the Bezold-Jarisch reflex: bradycardia, vasodilation, and hypotension. ^[11]^ Excessive vagus nerve excitation is an important factor that may cause intra-procedural hypotension and bradycardia. We should inhibit excessive vagus nerve excitation to prevent and treat intra-procedural hypotension and bradycardia.

Using thrombus aspiration devices during operation is more likely to evoke intra-procedural hypotension based on our research.Patients with using thrombus aspiration devices during operation were at 1.793 higher risk of intra-procedural hypotension than patients without using thrombus aspiration devices in our study.

Complete occlusion of culprit vessel is a independent risk factor of intra-procedural hypotension. The good patency of infarct-related arteries before PPCI indicates that the thrombus burden is lighter, spontaneous endogenous thrombolysis and the release of vasospasm.^[12]^In our study, patients with completely occlusion of culprit vessel were at 4.35 higher risk of intra-procedural hypotension than patients without completely occlusion of culprit vessel.

Our diagnostic model of intra-procedural hypotension was not a relative value but an absolute value. It was developed in an unselected real-world population. The nomogram we constructed for intra-procedural hypotension captures the majority of diagnostic information offered by a full logistic regression model and was more readily used at the bedside.

### Study Limitations

This a retrospective unicenter registry. Some patients were enrolled >10 years ago thus their treatment may not conform to current standards and techniques. No-reflow would not alter the interventionalist’s approach too much who is already “in” the situation of re-establishing flow into the vessel. The c statistic of the study intra-procedural hypotension model at 0.685 in the derivation and 0.718 in the validation cohort was modest.

## Conclusions

We developed and externally validated a moderate diagnostic model of intra-procedural hypotension during PPCI. We can use the formula or nomogram to predict intra-procedural hypotension.

## Data Availability

Data Availability Statement

The data used to support the findings of this study are included within the supplementary information file.

Supplementary Materials

The data are demographic, and clinical characteristics of hospitalized patients with acute STEMI. AGE = age; COCV=complete occlusion of culprit vessel; CNR = coronary artery no-reflow phenomenon; HCAD = history of of coronary artery disease; HH = history of hypertension; HD = history of diabetes; IH = intra-procedural hypotension; LAD = the culprit vessel was left anterior descending coronary artery; LCX = the culprit vessel was left circumflex coronary artery; RCA = the culprit vessel was right coronary artery; S = sex; TA= using thrombus aspiration devices during operation.

https://pan.baidu.com/s/1toHX7u3IKRgUfI2Bt4KmBQ

## Abbreviations

AMI: Acute myocardial infarction
AUC: Area under the receiver operating characteristic curve
LAD: Left anterior descending.
LCX: Left circumflex coronary artery
MI: Myocardial infarction
PCI: Percutaneous coronary intervention
PPCI: Primary percutaneous coronary intervention
RCA: Right coronary artery
ROC: Receiver operating characteristic
SBP: Systolic blood pressure
STEMI: ST elevation myocardial infarction
TIMI: Trombolysis in myocardial infarction flow grade

## Ethical Approval

The Ethics Committee of Beijing Anzhen Hospital, Capital Medical University, approved the study (No:2019032X). This study was registered with WHO International Clinical Trials Registry Platform (ICTRP) on 6 September 2019 (No: ChiCTR1900025706). http://www.chictr.org.cn/edit.aspx?pid=42913&htm=4.

## Statement of human and animal rights

All procedures performed in studies involving human participants were in accordance with the ethical standards of the institutional and/or national research committee and with the 1964 Helsinki declaration and its later amendments or comparable ethical standards. The study was not conducted with animals.

## Contributorship Statement

Yong Li contributed to generating the study data, analysed, interpreted the study data, drafted the manuscript, and revised the manuscript. Yong Li is being responsible for the overall content as guarantor. All authors have read and approved the manuscript.

## Acknowledgments

None.

## References

[1] Azzalini L, Poletti E, Lombardo F, et al. Risk of contrast-induced nephropathy in patients undergoing complex percutaneous coronary intervention. Int J Cardiol 2019;290:59–63.

[2] Demir OM, Lombardo F, Poletti E, et al. Contrast-Induced Nephropathy After Percutaneous Coronary Intervention for Chronic Total Occlusion Versus Non-Occlusive Coronary Artery Disease. Am J Cardiol 2018;122:1837–42.

[3] Karabağ Y, Çağdaş M, Rencuzogullari I, et al. The C-Reactive Protein to Albumin Ratio Predicts Acute Kidney Injury in Patients With ST-Segment Elevation Myocardial Infarction Undergoing Primary Percutaneous Coronary Intervention. Heart Lung Circ. 2019. 28(11): 1638–1645.

[4] Moons KG, Altman DG, Reitsma JB, et al. Transparent Reporting of a multivariable prediction model for Individual Prognosis or Diagnosis (TRIPOD): explanation and elaboration. Ann Intern Med 2015;162:W1–73.

[5] Thygesen K, Alpert JS, Jaffe AS, et al. Fourth Universal Definition of Myocardial Infarction (2018). J Am Coll Cardiol, 2018,72(18):2231–2264.

[6] Baran DA, Grines CL, Bailey S, et al. SCAI clinical expert consensus statement on the classification of cardiogenic shock: This document was endorsed by the American College of Cardiology (ACC), the American Heart Association (AHA), the Society of Critical Care Medicine (SCCM), and the Society of Thoracic Surgeons (STS) in April 2019. Catheter Cardiovasc Interv. 2019. 94(1): 29–37.

[7] Rezkalla SH, Kloner RA. Coronary no-reflow phenomenon: from the experimental laboratory to the cardiac catheterization laboratory.Catheter Cardiovasc Interv. 2008;72:950–957.

[8] Rezkalla SH, Stankowski RV, Hanna J, Kloner RA. Management of No-Reflow Phenomenon in the Catheterization Laboratory. JACC Cardiovasc Interv 2017;10:215–23.

[9] Ustinova EE, Schultz HD. Activation of cardiac vagal afferents in ischemia and reperfusion. Prostaglandins versus oxygen-derived free radicals. Circ Res 1994;74:904–11.

[10] Li Y, Lyu S. Risk Factors of Periprocedural Bradycardia during Primary Percutaneous Coronary Intervention in Patients with Acute ST-Elevation Myocardial Infarction. Cardiol Res Pract. 2019. 2019: 4184702.

[11] Campagna JA, Carter C. Clinical relevance of the Bezold-Jarisch reflex[J]. Anesthesiology, 2003,98(5):1250–1260.

[12] Kirma C, Izgi A, Dundar C, et al. Clinical and procedural predictors of no-reflow phenomenon after primary percutaneous coronary interventions: experience at a single center. Circ J 2008;72:716–21.

[13] Syed T, Tamis-Holland J, Coven D, Hong MK. Can glycopyrrolate replace temporary pacemaker and atropine in patients at high risk for symptomatic bradycardia undergoing AngioJet mechanical thrombectomy. J Invasive Cardiol 2008;20:19A–21A.

